# The rs11150601 intron variant of SETD1A is associated with female schizophrenia in the UK Biobank cohort

**DOI:** 10.1101/2024.12.01.24318271

**Authors:** Steven Lehrer, Peter H. Rheinstein

## Abstract

**Background:** Schizophrenia is a complex psychiatric disorder with an estimated heritability of 80%. SETD1A, a gene encoding a histone methyltransferase critical for transcriptional regulation, has been identified as a significant risk factor for schizophrenia. Loss-of-function mutations in SETD1A confer up to a 35-fold increased risk, implicating its role in neurodevelopment and synaptic plasticity.

**Methods:** Using data from the UK Biobank cohort (468,998 participants), we investigated the association of SETD1A variants with schizophrenia, obesity, and hypertension. Schizophrenia cases were identified using ICD-10 codes, while obesity and hypertension were assessed using specific data fields. Genome-wide association analysis was performed using PLINK, and statistical analyses utilized SPSS v26. Logistic regression assessed the impact of SETD1A intron variant (rs11150601) alongside age, obesity, and hypertension on schizophrenia risk.

**Results:** Among 1,063 individuals diagnosed with schizophrenia, obesity (p < 0.001) and hypertension (p < 0.001) were significantly more prevalent. The rs11150601 GG genotype was associated with an increased risk of schizophrenia in women (OR 1.6, p < 0.001) but not in men. Logistic regression revealed that obesity, hypertension, and age were independent risk factors for schizophrenia in women. SETD1A genotype exerted a significant sex-specific effect, highlighting its potential role in the biological mechanisms underlying schizophrenia.

**Conclusion:** Our findings emphasize the role of SETD1A in the genetic architecture of schizophrenia and its comorbidities, particularly in women. The sex-specific effects of SETD1A variants underscore the importance of incorporating biological sex into studies of psychiatric genetics. Further research is warranted to elucidate the mechanisms by which SETD1A influences neurodevelopment and identify therapeutic strategies targeting its epigenetic functions.

**Graphical abstract:** The rs11150601 intron variant of SETD1A is associated with female schizophrenia but not male schizophrenia.

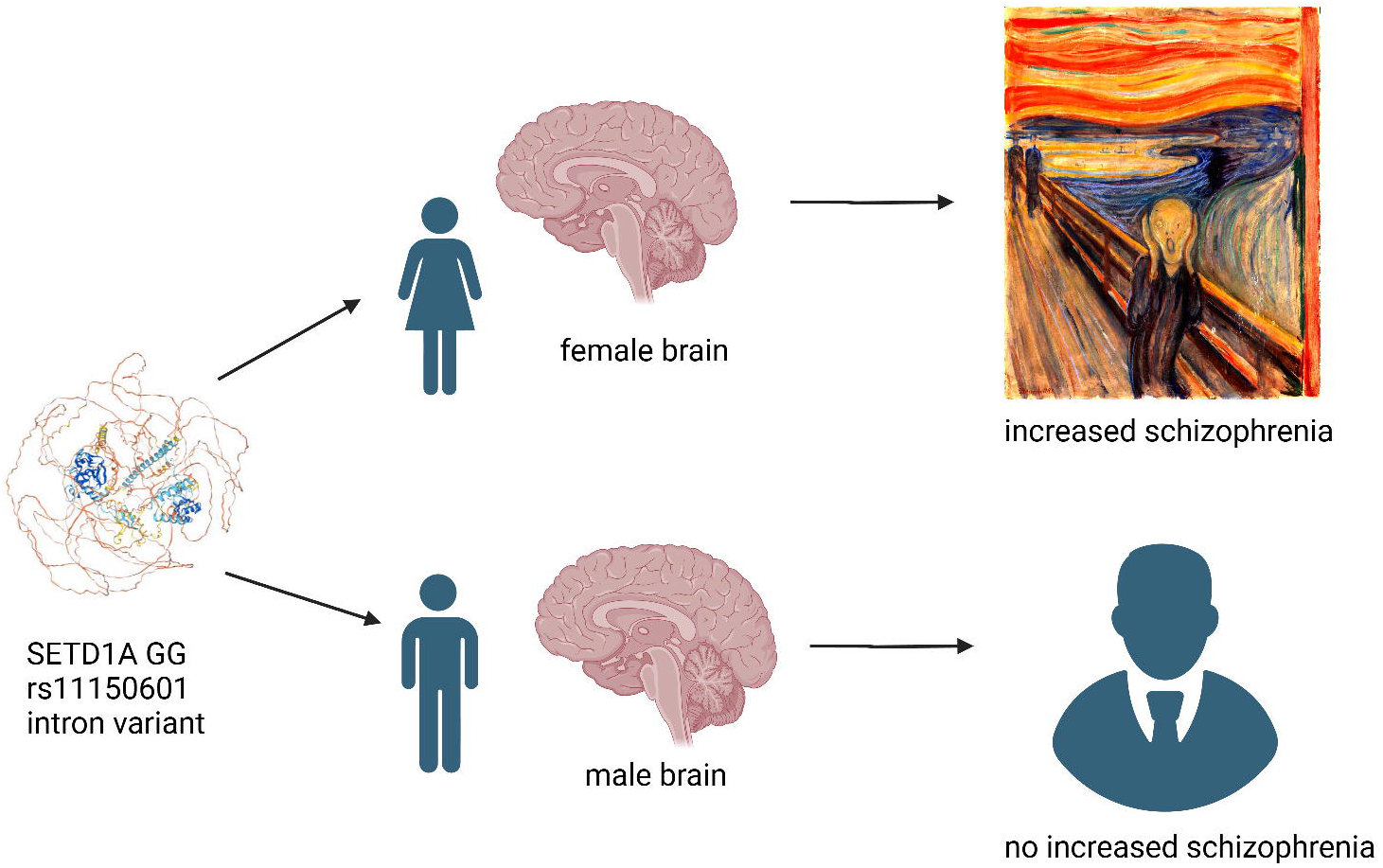

---

Schizophrenia is a chronic, severe mental disorder that affects how a person thinks, feels, and behaves. People with schizophrenia may seem like they have lost touch with reality. The exact cause of schizophrenia is unknown; the disorder probably results from a combination of genetic, brain chemistry, and environmental factors.

Schizophrenia has a strong genetic component, with heritability estimates around 80%. Unlike single-gene disorders, schizophrenia is polygenic. Over 100 genetic loci have been associated with an increased risk of schizophrenia.

Two major studies led by the Broad Institute identified genes and genomic regions linked to schizophrenia risk. The research, involving large international cohorts, pinpointed 287 genetic loci, highlighting pathways critical to brain development and synaptic function [1, 2].

SETD1A (SET domain containing protein 1A), a key histone methyltransferase involved in chromatin remodeling and gene expression regulation, emerged as a significant genetic risk factor for schizophrenia. Rare loss-of-function mutations in *SETD1A* were strongly associated with a marked increase in schizophrenia risk, implicating its role in the neurodevelopmental processes underlying the disorder. Variants in the SETD1A gene, particularly loss-of-function mutations, raised the risk by up to 35-fold. This makes SETD1A one of the most important genetic factors in schizophrenia. [3].

In the current study we used PheWeb and UK Biobank (UKB) data to further elucidate the role of SETD1A in schizophrenia, especially as it relates to obesity and hypertension, which are strongly associated with schizophrenia [4-6]. We included the rs11150601 intron variant of SETD1A that has been associated with Parkinson’s disease [7].

## Methods

PheWeb is a web-based tool designed for visualizing and browsing through genetic association results. It allows users to interactively explore genome-wide association studies (GWAS) and phenome-wide association studies (PheWAS) results. PheWeb is often used to analyze genetic data from large biobanks, such as the UK Biobank and the Michigan Genomics Initiative (MGI).

The UK Biobank is a large-scale biomedical database and research resource containing detailed genetic, lifestyle, and health information from 500,000 UK participants. Established in 2006, it aims to improve the prevention, diagnosis, and treatment of a wide range of serious illnesses, including cancer, heart disease, stroke, diabetes, and dementia.

Cases of schizophrenia were identified in UKB using ICD-10 classification F20. Cases of obesity were identified with UKB Data-Field 130792, date of first reported obesity. Cases of hypertension were identified with UKB Data-Field 131286, date first reported essential (primary) hypertension.

Whole genome association analysis was performed with PLINK and PLINK2.

Statistical analysis was done with SPSS v26 (IBM, New York).

## Results

Records of 468,998 subjects were examined. 94% were White British. Age was 56 ± 8 (mean ± SD). 1,063 subjects were schizophrenic. The rs11150601 intron variant of SETD1A, 16p11.2 A > G, has minor allele (G) frequency 0.38 in UKB.

Table 1 shows phewas results for SETD1A, 10799 cases, 397993 controls. Obesity and hypertension were the phenotypes with the strongest associations.

**Table 1.** Phewas results SETD1A. 10799 cases, 397993 controls. Obesity and hypertension were the phenotypes with the strongest associations.

| Top p-value in gene | Phenotype |
| --- | --- |
| 2.90E-08 | Obesity |
| 6.20E-08 | Overweight, obesity and other hyperalimentation |
| 6.90E-08 | Hypertension |
| 7.60E-08 | Essential hypertension |

**Table 1.** Schizophrenia versus SETD1A genotype in women and men. Significantly more women schizophrenics versus non-schizophrenics were GG homozygotes, 22.4% versus 15.3% (p < 0.001, two tailed Fisher exact test). SETD1A genotype had no significant effect on schizophrenia in men, GG homozygotes 15.3% versus 14.9% (p = 0.961).

|  |  |  | Schizophrenia |  |  |  |
| --- | --- | --- | --- | --- | --- | --- |
|  | Genotype |  | No | Yes | Total |  |
| Women | AA | Count | 101169 | 135 | 101304 | p < 0.001 |
|  |  | % within schizophrenia | 38.40% | 33.20% | 38.40% |  |
|  | AG | Count | 122036 | 181 | 122217 |  |
|  |  | % within schizophrenia | 46.30% | 44.50% | 46.30% |  |
|  | GG | Count | 40433 | 91 | 40524 |  |
|  |  | % within schizophrenia | **15.30%** | **22.40%** | 15.30% |  |
| Men | AA | Count | 85406 | 255 | 85661 | p = 0.961 |
|  |  | % within schizophrenia | 38.40% | 38.90% | 38.40% |  |
|  | AG | Count | 102865 | 303 | 103168 |  |
|  |  | % within schizophrenia | **46.30%** | **46.20%** | 46.30% |  |
|  | GG | Count | 34026 | 98 | 34124 |  |
|  |  | % within schizophrenia | 15.30% | 14.90% | 15.30% |  |

Figure 1A shows obesity and schizophrenia in subjects analyzed. Obesity was significantly increased in schizophrenics (p < 0.001, two tail Fisher exact test). Figure 1B shows hypertension in subjects analyzed. Hypertension was significantly increased in schizophrenics (p < 0.001, two tail Fisher exact test).

**Figure 1.**
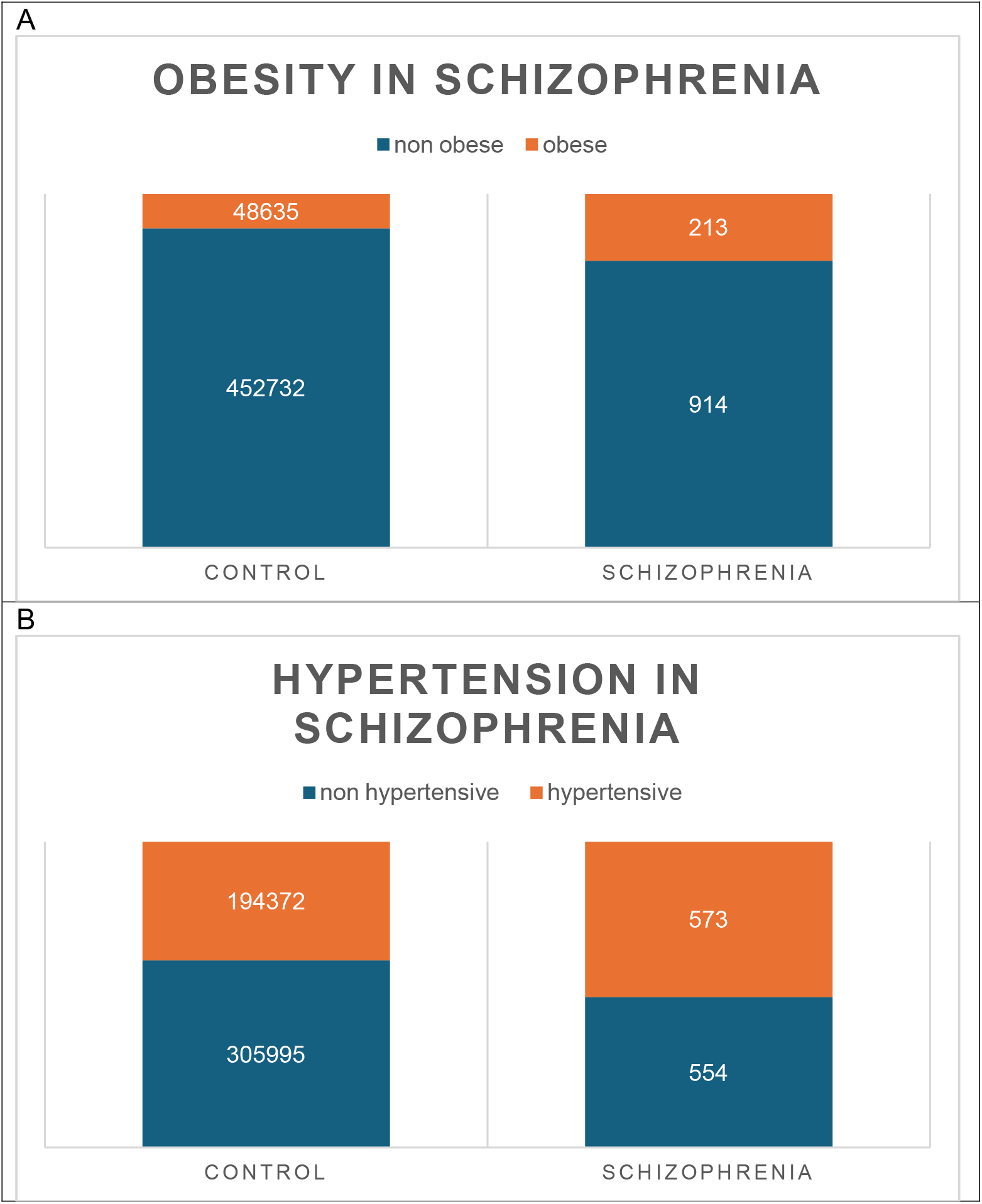
A. Obesity and schizophrenia in subjects analyzed. Obesity was significantly increased in schizophrenics (p < 0.001, two tail Fisher exact test). Figure 1B. Hypertension and schizophrenia in subjects analyzed. Hypertension was significantly increased in schizophrenics (p < 0.001, two tail Fisher exact test).

Table 2 shows schizophrenia versus SETD1A genotype in women and men. Significantly more women schizophrenics than non-schizophrenics were GG homozygotes, 22.4% versus 15.3% (p < 0.001, two tailed Fisher exact test). SETD1A genotype had no significant effect on schizophrenia in men, GG homozygotes 15.3% non-schizophrenic versus 14.9% schizophrenic (p = 0.961).

**Table 2.** Logistic regression, schizophrenia (yes, no) in women dependent variable, independent variables age, obesity (yes, no), hypertension (yes, no), SETD1A genotype, AG versus AA, GG versus AA. Risk of schizophrenia increased for each year decrease of age (odds ratio 0.974, p < 0.001). Risk of schizophrenia increased with obesity (O.R. 2.56, p < 0.001). Risk of schizophrenia increased with hypertension (O.R. 2.024, p < 0.001). Women heterozygous for the minor allele AG were at insignificantly increased risk of schizophrenia (O.R. 1.1, p = 0.403). Women homozygous for the minor allele GG were at significantly increased risk of schizophrenia (O.R. 1.6, p < 0.001). L.B. lower bound, U.B. upper bound.

|  | 95% L.B. | O.R. | 95% U.B. | p value |
| --- | --- | --- | --- | --- |
| age | 0.963 | 0.974 | 0.986 | <0.001 |
| obesity | 2.027 | 2.560 | 3.232 | <0.001 |
| hypertension | 1.638 | 2.024 | 2.501 | <0.001 |
| AG vs AA | 0.879 | 1.100 | 1.375 | 0.403 |
| GG vs AA | 1.226 | 1.600 | 2.089 | <0.001 |

Table 3 shows logistic regression, schizophrenia (yes, no) in women dependent variable, independent variables age, obesity (yes, no), hypertension (yes, no), SETD1A genotype, AG versus AA, GG versus AA. Risk of schizophrenia increased for each year decrease of age (odds ratio 0.974, p < 0.001). Risk of schizophrenia increased with obesity (O.R. 2.56, p < 0.001). Risk of schizophrenia increased with hypertension (O.R. 2.024, p < 0.001). Women heterozygous for the SETD1A minor allele AG were at insignificantly increased risk of schizophrenia (O.R. 1.1, p = 0.403). Women homozygous for the SETD1A minor allele GG were at significantly increased risk of schizophrenia (O.R. 1.6, p < 0.001).

## Discussion

SETD1A mediates the methylation of H3K4, a hallmark of active transcription. Aberrant H3K4 methylation alters the expression of genes essential for synaptic plasticity, such as NRXN1 and SHANK3, contributing to disrupted connectivity in neural networks [8]. By modifying chromatin structure, SETD1A enables the precise regulation of genes essential for neural differentiation, synaptic development, and plasticity.

During brain development, SETD1A controls the expression of genes involved in neuronal migration, axonal guidance, and synapse formation. Disruption of these processes due to SETD1A mutations can impair neural circuitry, potentially contributing to the structural and functional brain abnormalities observed in schizophrenia. SETD1A mutations are associated with reduced cortical thickness and impaired prefrontal cortex function, a hallmark of schizophrenia.

Genome-wide studies and exome sequencing have linked rare loss-of-function mutations in SETD1A to schizophrenia, with carriers showing up to a 35-fold increased risk [3].

These mutations also appear in related neuropsychiatric disorders, including intellectual disability and autism spectrum disorders, suggesting shared mechanisms of neurodevelopmental disruption.

Haploinsufficiency is a genetic condition that occurs when one copy of a gene is deleted or inactivated, and the remaining copy is unable to produce enough of the gene product to maintain normal function. SETD1A haploinsufficient mice exhibit behavioral deficits analogous to schizophrenia, including impaired working memory, social interaction, and sensorimotor gating. These phenotypes are accompanied by disruptions in synaptic morphology and function. Induced pluripotent stem cell (iPSC)-derived neurons from individuals with SETD1A mutations show altered dendritic arborization, reduced synaptic density, and aberrant electrophysiological activity. These findings underscore SETD1A’s role in synaptic maturation and stability [9-11].

The SETD1A intron variant rs11150601 is strongly association with both Alzheimer’s disease and related dementias (ADRD) and Parkinson’s disease (PD). The variant increases the risk of PD (odds ratio [OR] = 1.09, p = 5.1 × 10^−2^□) and has a lesser effect on ADRD (OR = 1.03, p = 1.5 × 10^−^□). These findings emphasize the shared genetic pathways influencing many neurodegenerative disorders [7].

The rs11150601 intron variant of SETD1A is not the first genetic variant in schizophrenia affecting women. The reelin gene (RELN) has been found to increase the risk of schizophrenia specifically in women. This variant is estimated to augment the risk by 1.4-fold for women carrying it. The reelin gene is known to play a role in brain development, but the exact mechanisms by which this variant predisposes women to schizophrenia are still unclear [12].

RELN and SETD1A are not directly related genes, but they both play important roles in neurodevelopment and brain function.

### Our study has weaknesses

Individuals with schizophrenia in the UK Biobank are those less severely affected [13].

The genetics of schizophrenia can vary by race, and our UK Biobank subjects were 94% white British. Certain genetic variants associated with schizophrenia may be more prevalent in specific racial or ethnic groups. For example, some genetic studies have found that individuals of African descent may have different genetic risk factors for schizophrenia compared to those of European or Asian descent [14, 15].

In conclusion, understanding SETD1A’s regulatory networks offers potential therapeutic avenues. Pharmacological agents that modulate epigenetic marks, such as histone deacetylase (HDAC) inhibitors, could potentially restore transcriptional balance. Emerging CRISPR-based tools could correct SETD1A mutations, offering a long-term solution to mitigating the genetic risks of schizophrenia.

## Data Availability

Data available from UK Biobank after approved application; data available from Pheweb without application

https://www.ukbiobank.ac.uk/

https://pheweb.org/UKB-SAIGE/about

